# Therapeutic Efficacy of Artemether-Lumefantrine (AL) plus Single Low Dose Primaquine for the treatment of uncomplicated *falciparum* malaria in a high transmission setting, Western Ethiopia

**DOI:** 10.1101/2025.10.19.25338327

**Authors:** Jimma Dinsa Deressa, Sinknesh Wolde Behaksra, Eshetu Molla, Girma Shumie, Bethlehem Adnew, Dawit Hailu Alemayehu, Fikregabrail Aberra Kassa, Alemayehu Letebo, Tamrayehu Seyoum, Elias B. Tafa, Kidist Woldekidan, Legesse Alamerie Ejigu, Tiffany Huwe, Migbaru Keffale, Gudissa Aseffa, Cristian Koepfli, Bayissa Chala, Yehenew Asmamaw, Fitsum Girma Tadesse, Endalamaw Gadisa

## Abstract

**Background:** The development and spread of drug-resistant parasites continue to threaten the move toward malaria elimination. Therapeutic efficacy and resistant marker studies are needed to guide national control programs. In African settings, evidence of partial resistance to artemisinin combination therapies associated with *Kelch13* is accumulating as World Health Organization (WHO) recommends a regular monitoring of first line antimalarial drugs to early detection of resistant parasites. In our study, we evaluated the efficacy of artemether-lumefantrine (AL) combined with a single low dose of primaquine (PQ) for treating uncomplicated *Plasmodium falciparum* malaria in a co-endemic area where *P. falciparum* is predominant.

**Methods and findings:** One hundred twenty-three cases with *P. falciparum* mono-infection were enrolled and treated with artemether-lumefantrine (AL) plus single low dose primaquine (PQ) as per the national guideline and followed up over 28 days. *Pfmsp2* capillary electrophoresis (CE) genotyping was used to differentiate recrudescence from new infection and we found seven recrudescence as true treatment failure. More than half (56.1%) of the participants had high (>10,000 parasites/μL) parasitemia at enrollment. At day 3, 17% (20/118) remained parasitemic and of the 10 individuals with detectable gametocytes at enrollment, only 1 remained gametocytemic on day 3, and 100% parasite clearance was achieved on day 7 respectively, indicating no early treatment failure. Multiplicity of infection was 3.8 and 1.7 before treatment and the day of recurrence respectively. The adequate clinical and parasitological responses, the PCR uncorrected at 28-day was 73.7% (95%_CI_ 65.6 - 82.9), while the PCR adjusted efficacy was 91.3% (95%_CI_ 85.3 – 97.7). In our study assessment, no serious adverse event was recorded as AL plus single low-dose PQ is safe for the treatment of uncomplicated falciparum malaria in the study setting.

**Conclusions:** The low efficacy found in our study is concerning, which might pos a challenge for malaria control in the study setting and beyond. We suggest that regular monitoring and conducting further research using advanced molecular techniques such as next-generation sequencing (NGS) to enable early detection of mutant variants that reduce treatment efficacy.

**Author Summary:** Since 2022, Ethiopia’s progress toward malaria elimination has been challenged by a nationwide surge in malaria cases, prompting intensified efforts by the national malaria control program to address this upsurge (Fig 1). Of the factors attributed to the widespread outbreak include the emergence and spread of diagnostic and drug resistance mutations. Due to the spread of the HRP2/3 deletion, the region is moving away from a rapid diagnostic test that has high sensitivity and specificity. On top, of the emergence and spread of the *Pfkelch13* mutation implicated in partial resistance to artemisinin-based combination, the first line regimen could be an impending challenge (Fig 2). The WHO recommends a regular therapeutic efficacy study to monitor antimalarial drugs with a target of 90% threshold for first line regimens.

Western Ethiopia has the highest malaria burden in the country. The Artemisinin Lumefantrine based combination is the first line for the treatment of uncomplicated falciparum malaria since the 2000s. In our study, we evaluated the efficacy of artemether-lumefantrine (AL) combined with a single low dose of primaquine (PQ) for treating uncomplicated *P. falciparum* malaria in a coendemic area where P. falciparum is predominant. A single arm partially directly observed 28days of follow up for one hundred twenty-three microscopically mono-infection were enrolled a therapeutic efficacy study, at Bambasi Health Center, Northwest Ethiopia. We found 91.3% PCR based adjusted efficacy (CE), a lower efficacy reported so far to the best of our knowledge in Ethiopia.

The PCR-corrected efficacy of 91.3%, just above the WHO’s 90% threshold, raises concerns about the declining effectiveness of artemether-lumefantrine (AL) in this high-transmission setting. We observed a high multiplicity of infection (MOI), with 98 Pfmsp2 allelic variants detected and an initial MOI of 3.8 at recruitment, which reduced to 1.7 on the day of recurrence, suggesting a complex parasite population that may contribute to treatment challenges. The WHO recommends amplicon-based sequencing in high MOI settings to distinguish recrudescence from new infections. Our findings, using Pfmsp2 CE genotyping, revealed a 28-day PCR-corrected efficacy of 91.3% for AL plus single low-dose PQ, which is below the 98.7% pooled efficacy reported in Ethiopian systematic reviews. Despite achieving 100% parasite clearance by day 7, the combination of AL and PQ showed enhanced gametocyte clearance compared to AL alone, though clearance was delayed, with one participant remaining gametocytemic on day 3. Pretreatment parasite density and hemoglobin levels likely influenced these outcomes. Notably, the high initial parasite load in 56.1% of participants (geometric mean of 13,513 parasites/μL) may have contributed to delayed clearance and elevated treatment tolerance risk. The efficacy of single low dose primaquine as WHO-recommended transmission blocking strategy requires further evaluation in a powered study, given the reduced efficacy observed in our study.

## Introduction

Malaria remains an important public health concern globally. As per the World Health Organization (WHO) estimate, 249 million malaria cases and 608,000 associated deaths were registered in 2022. Ethiopia is among the high burden countries. Of the total cases, 94% and 95% of new cases and deaths, respectively, were from the WHO African region [1]. Between 2000 and 2015, the wide implementation of integrated control interventions resulted in an 18% and 48% reduction in malaria incidence and mortality, respectively [2]. During the same period, 663 million malaria cases were averted in sub-Saharan Africa, of which 21% were due to ACTs [2]. Malaria is a major health and socioeconomic challenge with over 81 million Ethiopians living at risk of malaria according to the national malaria strategic plan 2024/25-2026/27 (Draft NMSP, MOH). Falciparum malaria remained the major cause of morbidity and mortality though *P. vivax* co-exists in different proportions in all malarious areas of the country [3].

Ethiopia’s over five decades of malaria control program achieved a substantial decrease in related mortality and morbidity until 2018. Yet, the gains were eroded and the country witnessed a surge in cases through 2019-23 (Fig 1). The advent of the COVID-19 pandemic, *hrp2*/*3* gene deletions, internal conflicts, and invasion of *An. stephensi* were some of the attributed causes [3].

The detection of gene variants implicated in the therapeutic efficacy of artemisinin combination therapy (ACT)-based treatment called for increased vigilance in African [4]. Studies carried out in Rwanda and Uganda revealed mutations in the *kelch13* region, *R561H,* and *A675V,* linked to delayed parasite clearance [5, 6]. Recrudescence following AL treatment linked to wildtype *Pfcrt K76T* and *Pfmdr1 N86Y* gene variants were reported in Kenya, Tanzania, and Zanzibar [7–10]. The K13 mutation implicated in AL partial resistance in Ethiopia, Sudan, and Eritrea showed an increased detection rate over time [11, 12]. In a *P. falciparum* and *P. vivax* co-endemic setting, in samples from AL plus a single low-dose PQ therapeutic efficacy trial, we genotyped the samples via capillary gel electrophoresis by targeting allelic variants of msp2 of FC27 and 3D7 to quantify multiplicity (complexity) of infection and differentiate the recrudescence from new infection.

## Methods

### Study area and period

This study was conducted at Bambasi health center (9°45′N 34°44′E), Bambasi district (Fig 3) from November 2020 to March 2021. *P. falciparum* was the dominant parasite species in the study setting [13]. The peak transmission season in the area is from September to December, and *An. gambiae sensu stricto* (and presumably *An. arabiensis*) is the primary vector [14].

**Figure 1:**
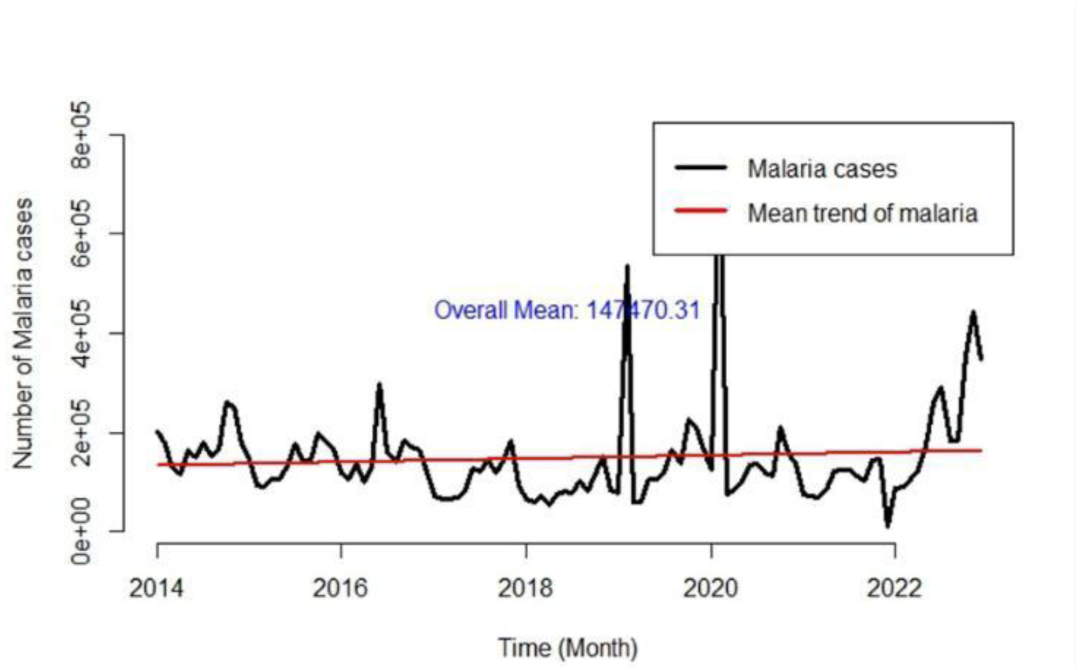
Trends of confirmed malaria cases in Ethiopia from 2014 to 2022.

**Figure 2:**
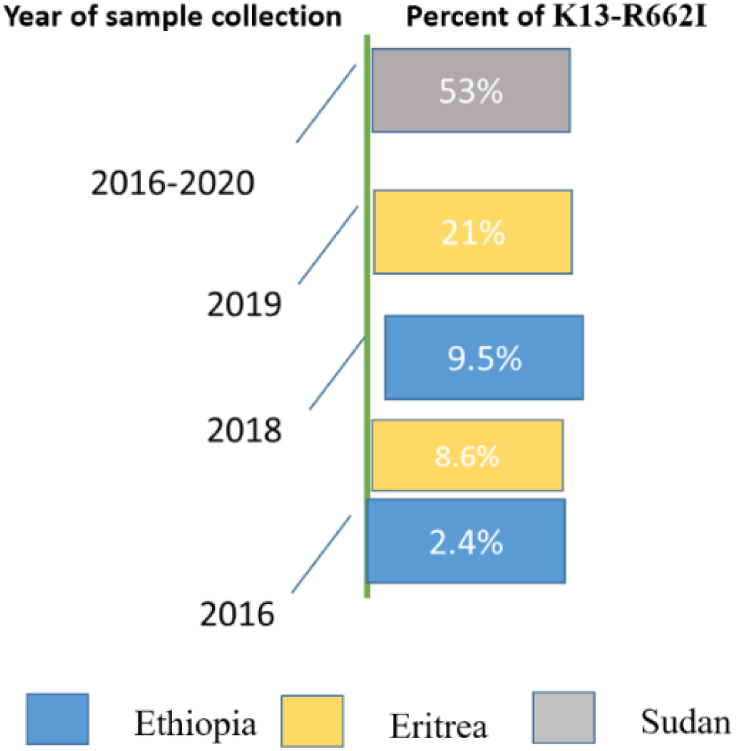
Ther change in PtK13-R6621 mutation.

**Fig 3.**
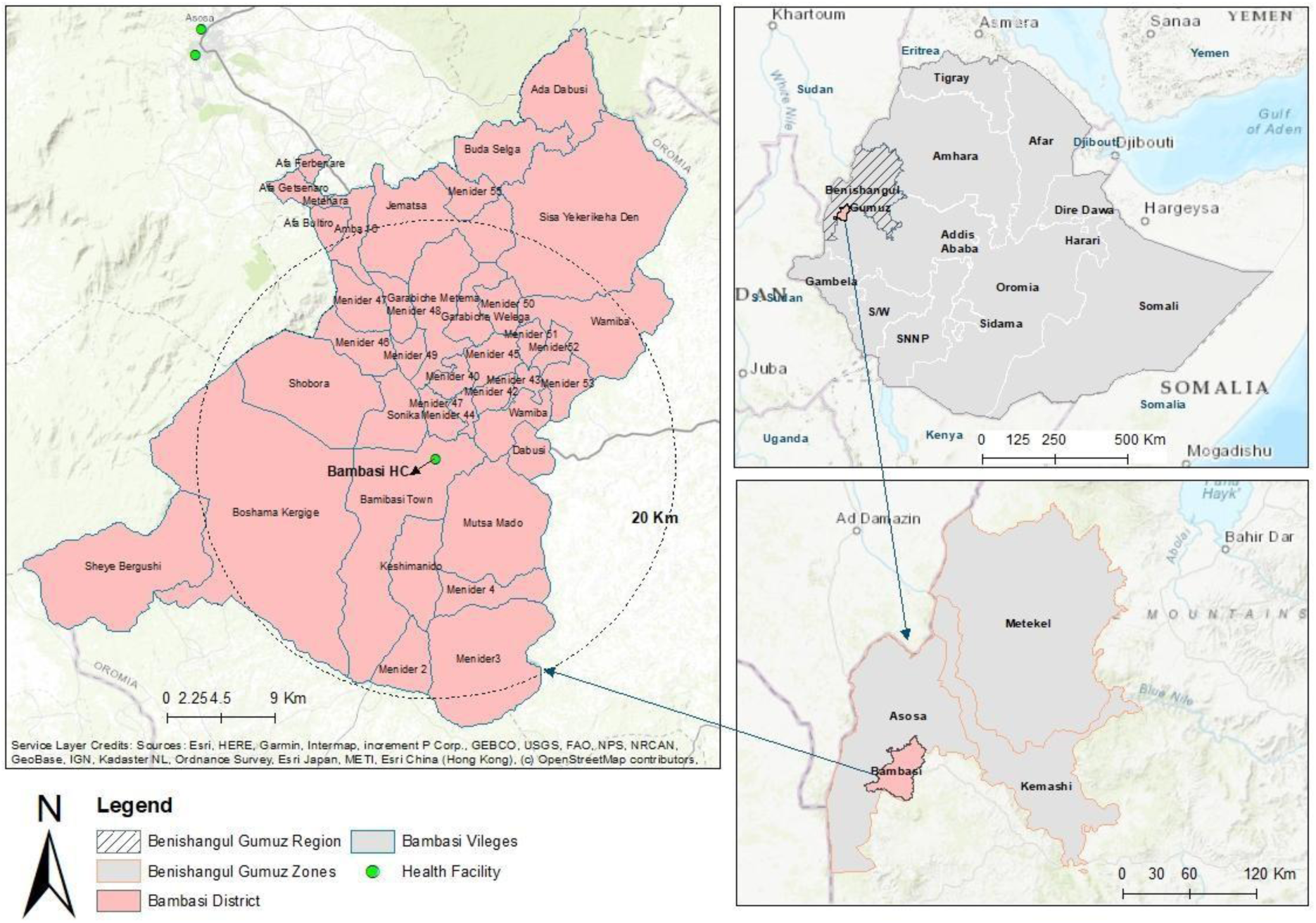
Map of the study site, Bambasi health center, Bambasi district, Benishangul Gumuz regional state, Northwest Ethiopia.

### Study design and Recruitment

A prospective single arm partially observed study was done following the methods for surveillance of antimalarial drug efficacy by WHO [15]. The sample size was determine based on the WHO drug efficacy evaluation criteria. Self-reporting febrile patients (axillary temperature ≥ 37.5°C) or with a history of fever within the past 24 hours at the time of diagnosis and those fulfilling the inclusion criteria were recruited. Microscopy and RDT confirmed uncomplicated *P. falciparum* mono-infected, 6 months or older, body weights >5kg, hemoglobin 5.0 g/dL and above, parasitemia ≥ 500 parasites/μL, HCG negative and/or non-breastfeeding, and residence within 20 km radius of Bambasi health center were included. Written informed consent or assent for those between the ages of 11 to 18 in addition to parental consent were sought. Clinical, and sociodemographic data were collected by clinical nurses/public health officers trained and research team members, and then, the data was stored on project data capturing tools (Redcap) with the data access to the contributors of this work.

### Drug regimens and follow-up

The study followed the national guideline to treat cases [16] and additionally, this in vivo efficacy trial study was registered under Pan-African clinical trials registry (PACTR) with unique identification number of PACTR202509595696440. Artemether-Lumefantrine (AL, 20 mg of Artemether and 120 mg of Lumefantrine; Novartis Pharmaceutical Corporation, New York, NY, USA) was administered twice daily for three days. The morning AL doses were directly observed, while the evening dose was taken at home and patients were asked if they appropriately took the drug as instructed by the study clinicians before administering the next dose and blisters were counted for doses taken at home to monitor adherence. The single dose of primaquine (PQ) (0.25mg/kg) was given on the day of enrollment 30 minutes after the first dose of AL. Participants were given vanilla cream biscuit (6*125g) with 0.5L bottled water while taking drugs and were advised to take the evening doses after a meal. Participants were observed for 30 minutes and replaced with the first dose in case of vomiting. An appointment card was given to each participant to come to the health center on days 1, 2, 3, 7, 14, 21, and 28 after dosing or at any unscheduled time when they had any of the malaria symptoms and/or hemolysis (darkening of their urine). The appointment card detailed the visit dates and pictorial guide of hemolysis. Further, they were communicated by phone a day earlier to the scheduled date; those who did not show up on schedule were visited at their residence for appropriate drug administration.

### Withdrawal and loss to follow-up

Participants were excluded from the study if they experienced a second vomiting during the first dose of AL, took self-prescribed antimalarial drugs, missed doses of AL, or were diagnosed with other than falciparum. Moreover, participants who couldn’t attend the health center on the scheduled date and couldn’t be reached by the research team/tracer within 1 or 2 days after the scheduled visit date were considered as a loss to follow-up. Participants who came with mixed infections or/and *P. vivax* infection at follow up were treated as per the national guideline [16], and excluded from the study.

### Laboratory procedures

Blood samples were collected from finger-prick for multi species CareStart rapid diagnostic test (RDT) (American Access Bio International Company) [17], thick and thin blood smears, hemoglobin assessment, and dried blood spot (DBS) for molecular analysis on days 0, 1, 2, 3, 7, 14, 21, and 28, and any unscheduled visit(s).

The thin smear was stained with 3% Giemsa and fixed in absolute methanol for 30 to 45 minutes and used for parasite speciation and quantification. Asexual parasite density per microliter was determined based on the number of parasites counted per 200 WBCs on a thick blood film. Two microscopists read the slides, and results with discordant by ≥400 parasites/µL were re-read by a third microscopist, blinded to the two results, and parasite density was calculated by averaging the two closest counts. Hemoglobin concentration was measured using a portable spectrophotometer (HemoCue®, Angelholm, Sweden), on days 0, 3, 14, and 28.

Genomic DNA was isolated from DBS using the Chelex method as described elsewhere [18, 19]. The amplification of *msp2* targeting FC27 and 3D7 allele loci was conducted on paired samples. The purified PCR products were genotyped following the protocol (Applied Biosystems, Foster City, CA, USA) and analyzed using capillary electrophoresis (Macrogen, Inc., Korea) [20]. Then, the capillary electrophoresis (CE) sequence analysis was performed using GeneMarker®V2.7.0 software (State College, PA) for allele sizing and scoring by both trained experts of the University of Notre Dame and Armauer Hansen Research Institute (AHRI).

### Treatment outcomes

The primary outcomes of this study were to determine the clinical and parasitological response of patients to antimalarial treatment with the proportion of patients who achieve the complete parasite clearance and resolution of symptoms. The treatment outcome was classified as Early Treatment Failure (ETF), Late Clinical Failure (LCF), Late Parasitological Failure (LPF), and Adequate Clinical and Parasitological Response (ACPR) [21]. The *Pfmsp2* genotyping was conducted to distinguish between new infections and recrudescence. The *msp2* amplicons were resolved through capillary electrophoresis (CE), allelic variants were identified and 3 base pairs or more differences were considered as new infections. Recurrent parasites are potentially classified into four based on the degree of allelic matching as earlier described in the study conducted by Mugittu et al 2006. These are; i) all alleles are identical at both baseline and recurrent parasite, ii) some alleles are missing in the recurrent parasites, iii) recurrent parasites contain alleles identical to those at baseline with additional/new ones not observed at baseline, and iv) all alleles in the baseline and recurrent parasites samples are different. It is generally accepted that categories from i-iii are represented as recrudescent while step four (iv) represents new infections [22].

### Data analysis

Data were entered into Microsoft Excel and imported into R-4.5.1version (Integrated Development for R Studio, PBC, Boston, USA) for statistical analysis. Descriptive statistics were used to calculate the frequency of MOI based on Pf*msp2* capillary gel electrophoresis genotyping. Chi square (χ2) tests were used to measure statistical significance (P<0.05) for specific outcomes, including differences in parasite clearance rates across age groups on follow-up days, and changes in anemia status between enrollment and follow-up days. Kaplan-Meier survival analysis was used to estimate the PCR-corrected and uncorrected ACPR at day 28.

## Results

### Participant characteristics

The mean age was 17.8 years (range 3 - 62), the majority of participants were male (61.0%), and most (57.7%) were in the 6 to 15 years age group. The geometric mean (GM) parasitemia was 13,513/μL (Table 1).

**Table 1.** Baseline characteristics of study participants stratified by age and sex.

| Age in years (n) | Sex (n) | Mean weight in kg (SD) | Auxiliary temp. in °C (SD) | Mean Hgb in g/dl (SD) | GM parasite/μL (95% CI) |
| --- | --- | --- | --- | --- | --- |
| **0.5 - 5(6)** | M (3) | 16 | 37.5 (0.8) | 10.3 (3.0) | 7209.8 (235.5 - 220700.9) |
|  | F (3) | 14 | 37.8 (0.7) | 12.6 (0.7) | 14364.6 (75.0 – 2751832.0) |
| **6 – 15 (71)** | M (44) | 32 | 37.3 (1.0) | 12.5 (1.8) | 14128.4 (10135.6 - 19694.1) |
|  | F (27) | 36 | 37.5 (1.0) | 13.3 (1.8) | 10891.4 (7068.3 - 16782.3) |
| **≥16 (46)** | M (28) | 56 | 37.5 (1.4) | 13.5 (1.7) | 17055.5 (9287.8 - 31319.7) |
|  | F (18) | 51 | 37.5 (1.6) | 12.7 (1.5) | 12815.6 (6092.9 - 26955.8) |
| **Total** |  | 40.4 | 37.5 (1.2) | 12.9 (1.8) | 13513.0 (10730.4 - 17017.1) |

SD = standard deviation, Kg = kilogram, Hb = haemoglobin, n = number, °C = degree centigrade, g/dl = gram/deciliter and μL= microliter, GM= Geometric Mean

### Asexual blood stage parasite, gametocyte, and fever clearance rate

Parasite clearance rates were 51.7% (61/118) and 83.0% (98/118) on day 2 and day 3 respectively. The decline in parasitemia was more pronounced (91.3%, 42/46) in the 16 years and above age groups. Of the 10 participants with microscopically detected gametocytes at enrollment, 5 and 1 remained gametocytemic on days 2 and 3 respectively. Most individuals (99.2%, 119/120) became afebrile 48 hours after dosing, and complete fever clearance was attained by day 3 (Table 2).

**Table 2.**
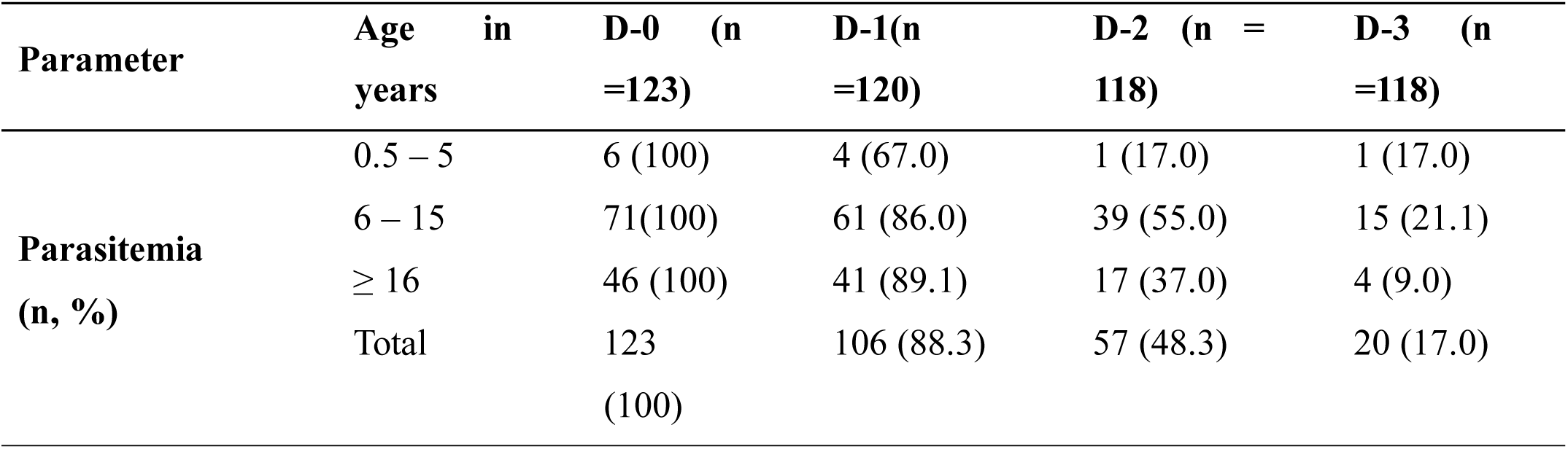
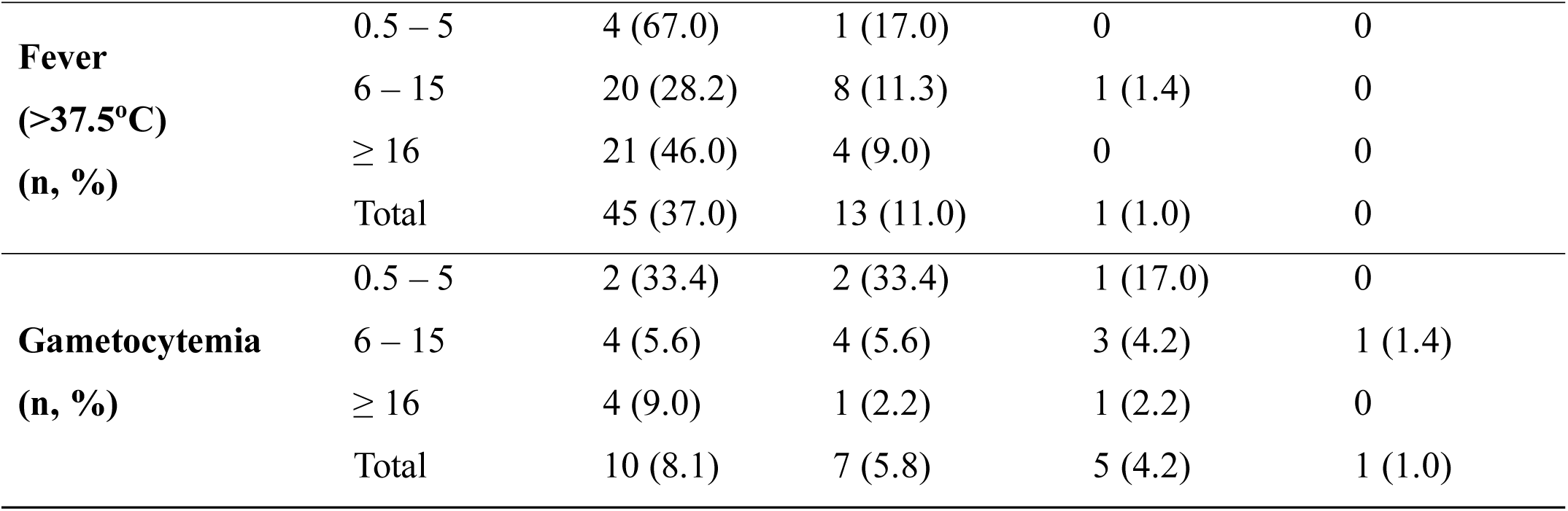
Parasite (by microscopy), and fever clearance rate in study participants Parameter.

| Parameter | Age in years | D-0 (n =123) | D-1(n =120) | D-2 (n = 118) | D-3 (n =118) |
| --- | --- | --- | --- | --- | --- |
| **Parasitemia (n, %)** | 0.5 – 5 | 6 (100) | 4 (67.0) | 1 (17.0) | 1 (17.0) |
|  | 6 – 15 | 71(100) | 61 (86.0) | 39 (55.0) | 15 (21.1) |
|  | ≥ 16 | 46 (100) | 41 (89.1) | 17 (37.0) | 4 (9.0) |
|  | Total | 123 (100) | 106 (88.3) | 57 (48.3) | 20 (17.0) |
| **Fever**<br>(**>37.5°C**)<br>(**n, %**) | 0.5 – 5 | 4 (67.0) | 1 (17.0) | 0 | 0 |
|  | 6 – 15 | 20 (28.2) | 8 (11.3) | 1 (1.4) | 0 |
|  | ≥ 16 | 21 (46.0) | 4 (9.0) | 0 | 0 |
|  | Total | 45 (37.0) | 13 (11.0) | 1 (1.0) | 0 |
| **Gametocytemia**<br>(**n, %**) | 0.5 – 5 | 2 (33.4) | 2 (33.4) | 1 (17.0) | 0 |
|  | 6 – 15 | 4 (5.6) | 4 (5.6) | 3 (4.2) | 1 (1.4) |
|  | ≥ 16 | 4 (9.0) | 1 (2.2) | 1 (2.2) | 0 |
|  | Total | 10 (8.1) | 7 (5.8) | 5 (4.2) | 1 (1.0) |

The proportion of anemia, as defined by the WHO Hb cut-off criteria [23], was 30% (37/123) on enrolment which declined to 20% at the end of the study (Table 3). There was no serious adverse event, the common adverse events reported were headache (35%), fever (21.8%), chills (14.7%), weakness (9.8%), nausea (6.5%), and abdominal pain (9.5%) respectively.

**Table 3.** Anemia status by age group on each follow-up day, Bambasi Health Center, Bambasi district, Northwestern Ethiopia, November 2020 to March 2021.

|  | Age category |  |  |  |
| --- | --- | --- | --- | --- |
|  | ≤5 (n, %) | 6-15 (n, %) | ≥16 (n, %) | Total (n, %) |
| **Anemia status at Do** |  |  |  |  |
| Anemic | 1, 16.7 | 19, 26.8 | 17, 37.0 | 37, 30.1 |
| Non-anemic | 5, 83.3 | 52, 73.2 | 29, 63.0 | 86, 69.9 |
| **Total** | **6, 100.0** | **71, 100.0** | **46, 100.0** | **123, 100.0** |
| **Anemia status at D3** |  |  |  |  |
| Anemic | 3, 60.0 | 30, 43.5 | 23, 52.3 | 56, 47.5 |
| Non-anemic | 2, 40.0 | 39, 56.5 | 21, 47.7 | 62, 52.5 |
| **Total** | **5, 100.0** | **69, 100.0** | **44, 100.0** | **118, 100.0** |
| **Anemia status at D14** |  |  |  |  |
| Anemic | 2, 40.0 | 13, 19.4 | 9, 20.9 | 24, 20.9 |
| Non-anemic | 3, 60.0 | 54, 80.6 | 34, 79.1 | 91, 79.1 |
| **Total** | **5, 100.0** | **67, 100.0** | **43, 100.0** | **115, 100.0** |
| **Anemia status at D28** |  |  |  |  |
| Anemic | 1, 33.3 | 4, 8.2 | 13, 34.2 | 18, 20.0 |
| Non-anemic | 2, 66.7 | 45, 91.8 | 25, 65.8 | 72, 80.0 |
| **Total** | **3, 100.0** | **49, 100.0** | **38, 100.0** | **90, 100.0** |

### Treatment outcomes

No early treatment failures (ETF) were observed. Of the 123 enrolled participants, only 73 completed the day-28 follow-up (Fig 4). The adequate clinical and parasitological responses at 28day (ACPR) was 73.7% (95% CI 65.6 - 82.9) and the PCR-corrected cure rate was 91.3% (95% CI 85.3 - 97.7) at day 28 (Table 4) and figure 5 respectively.

**Fig 4.**
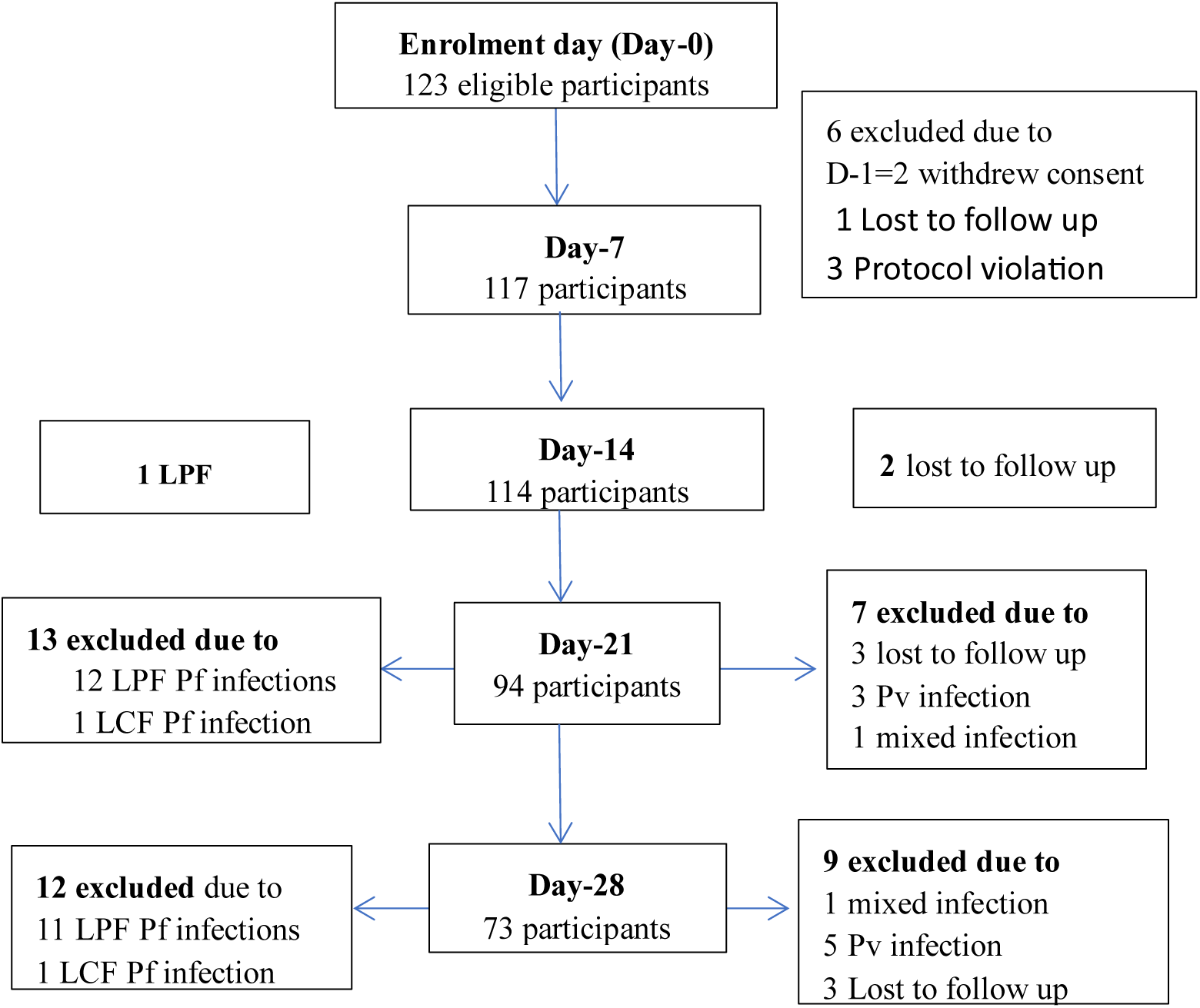
Details on study follow-up progress. Of the 123 eligible participants enrolled in this study, only 73 completed the 28 days of follow-up. LPF & LCF = late parasitological and clinical failure respectively.

**Fig 5.**
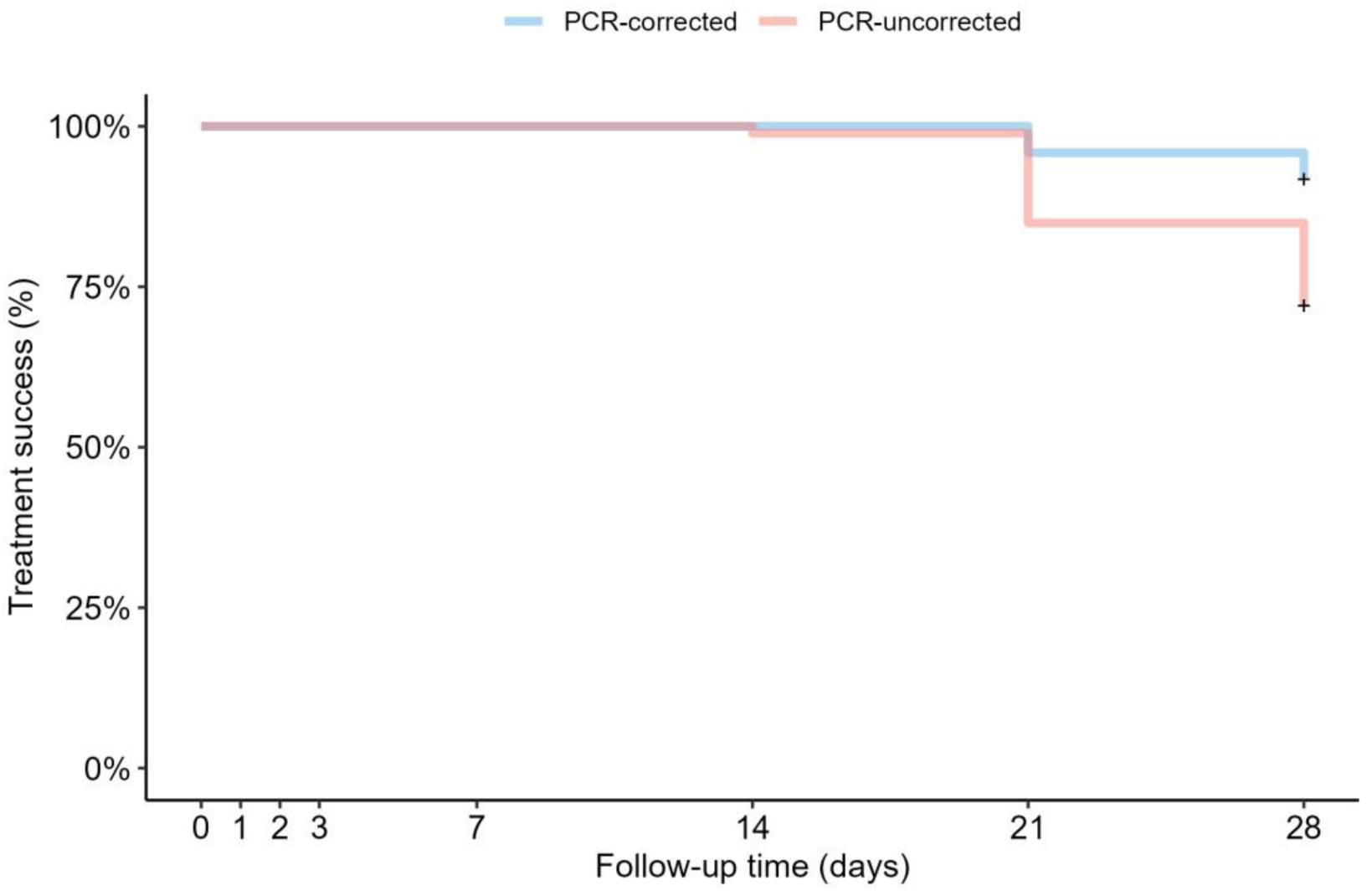
Kaplan-Meier analysis; CE-corrected and uncorrected cure rates for uncomplicated *P. falciparum* treated with AL plus single low-dose PQ, Bambasi, Northwest Ethiopia 2021. Kaplan-Meier analysis showed the PCR-corrected treatment success of 91.3% (95%_CI_ 85.3 – 97.7) at day 28 and 73.7% (95%_CI_ 65.6 - 82.9) at day 28 for PCR-uncorrected, respectively.

**Table 4.** Efficacy of AL plus single low-dose PQ for the treatment of uncomplicated *falciparum* malaria, Bambasi health center, Bambasi district, Northwest Ethiopia, November 2020 to March 2021.

| Treatment Outcome | Microscopy (PCR uncorrected) |  |  |  | *msp2* Capillary Electrophoresis (CE) (3bp) |  |  |  |
| --- | --- | --- | --- | --- | --- | --- | --- | --- |
|  | Age in Years, (n, %) |  |  | Total (n, %) | Age in Years, (n, %) |  |  | Total (n, %) |
|  | 0.5 - 5 (6) | 6 - 15 (71) | ≥16 (46) |  | 0.5 - 5 (6) | 6 - 15 (71) | ≥ 16 (46) |  |
| **ETF** | 0 (0.0) | 0 (0.0) | 0 (0.0) | 0 (0.0) | 0 (0.0) | 0 (0.0) | 0 (0.0) | 0 (0.0) |
| **LCF** | 1 (25.0) | 1 (1.8) | 0 (0.0) | 2 (2.1) | 1 (33.3) | 1 (2.3) | 0 (0.0) | 2 (2.5) |
| **LPF** | 1 (25.0) | 17 (29.8) | 6 (15.8) | 24 (24.2) | 0 (0.0) | 3 (7.0) | 2 (5.9) | 5 (6.3) |
| **ACPR** | 2 (50.0) | 39 (68.4) | 32 (84.2) | 73 (73.7) | 2 (66.7) | 39 (90.7) | 32 (94.1) | 73 (91.3) |
| **Analyzed** | 4 | 57 | 38 | 99 | 3 | 43 | 34 | 80 |
| **Withdrawal** | 1 | 2 | 1 | 4 | 2 | 22 | 8 | 32 |
| **LFU** | 1 | 6 | 3 | 10 | 1 | 6 | 3 | 10 |
| **Protocol violation** | 0 | 6 | 4 | 10 | 0 | 0 | 1 | 1 |
| **Total** | 6 | 71 | 46 | 123 | 6 | 71 | 46 | 123 |

Where: *Early treatment failure (ETF), late parasitological (LPF), and late clinical failure (LCF) respectively. and LFU; adequate clinical and parasitological response (ACPR) and loss to follow-up (LFU). Withdrawal; individuals with new infections and unidentified and protocol violations; individuals who withdrew their consent, parasite recurrent with vivax infections and/or mixed infections. Analyzed; the number of individuals eligible for analysis as per the WHO efficacy protocol.*

*Pfmsp2* CE genotyping successfully amplified in 26 paired samples, identifying 98 allelic variants at day 0 (52 FC27, 46 3D7) and 44 at recurrence (29 3D7, 15 FC27). The mean MOI was 3.8 at enrollment and 1.7 at recurrence. Seven true treatment failures (5 LPF, 2 LCF) were confirmed as recrudescence by CE genotyping.

## Discussion

Antimalarials remained at the top of the list of armaments in the malaria control and elimination endeavor. As the fight tightens and the diagnostic and appropriate treatment tools get biting the parasite also invests in ways of survival; developing resistance and spread. *Plasmodium falciparum* (*P. falciparum*) developed resistance to almost all antimalarials rolled out, thus surveillance strategies to continuously monitor such development have been put in place [24, 25]. The combination of *in vivo* therapeutic efficacy study and CE based molecular genotyping was employed in identifying recrudescence, is used in these efficacy monitoring studies. Here we present a study in which we evaluated AL plus single low-dose PQ for the treatment of uncomplicated *P. falciparum* malaria, implicated to assess the efficacy of the combination therapy. Our study site is a *P. vivax* and *P. falciparum* co-endemic area where *P. falciparum* is the predominant [13] with a high multiplicity of infection; we found 98 *msp2* allelic variants and an MOI of 3.8 at recruitment and 1.7 on the day of recurrence, respectively. We suggest that the observed decline in MOI post-treatment likely due to a combination of selective pressure exerted by the treatment, possibly leading to the survival of more resistant genotypes, and a reduction in genetic diversity as immune response and treatment clears susceptible strains. In areas with a high MOI, the WHO recommends amplicon-based sequencing [26] for identifying recrudescence, and combinations of *msp1*/*msp2*/*glurp* genotyping were used to distinguish new infections from recrudescence [27]. We used *msp2* capillary gel electrophoresis at a 3-base pair (bp) difference for the two allelic variants (3D7-FC27), and thus, efficacy results should be interpreted with caution. Our findings indicated a 28-day PCR-corrected efficacy of 91.3%, which is lower than previously reported efficacy rates for AL in Ethiopia and this suggests the potential challenges for malaria control and elimination program. Efficacy rates can vary significantly by regional variability due to local transmission intensity, parasite strains, nutritional status, co-morbidity, treatment adherence and severity of malaria during enrollment. Higher genetic diversity of the parasite can influence and lead to variations in susceptibility to AL, potentially, resulting lowering in overall efficacy as observed in our study conducted in high transmission setting with observed high complexity of infections. However, a systematic review and meta-analysis of 15 therapeutic efficacy studies that involved 1523 patients conducted from 2004 to 2020 documented a PCR-adjusted pooled success of 98.7% [28]. A more recent study done in two sentinel sites, western and southern Ethiopia, reported 100% treatment success after PCR adjustment [29]. A study done in Benishangul-Gumuz in 2020, close to our study site, reported 96% AL efficacy after PCR adjustment [30]. In our study, 88.3%, 48.3%, and 17% of participants had microscopy detectable parasitemia on day 1, 2 and day 3 respectively, while 100% clearance was achieved by day 7; residual submicroscopic and microscopic parasitemia raises concerns about treatment outcomes.

In the aforementioned study in Benishangul Gumuz [30], 38% and 10% of the participants remained microscopy positive on day 1 and day 2. Interestingly, 60% and 28% of participants had PCR detectable infection on days 3 and 7, respectively, while 20% of participants remained PCR positive at the last visit (day 28). In our study, AL+PQ reduced gametocyte carriage, with complete clearance by day 7, but prolonged clearance (1 participant gametocytemic on day 3) suggests limited transmission-blocking efficacy at the WHO-recommended PQ dose (0.25 mg/kg). This aligns with studies indicating that single low-dose PQ enhances gametocyte clearance but may not fully sterilize gametocytes [38]. Even though the smallest number of patients with detectable gametocyte observed during enrollment, the combination of AL and PQ demonstrated improved gametocyte clearance compared to AL alone, although the duration of gametocyte clearance was prolonged, with one participant remaining gametocytemic at day 3. Factors such as pretreatment parasite density and hemoglobin levels may have influenced these outcomes [31], however, this result showed a better gametocyte clearance rate compared to AL alone, treated delayed up to day 14, study conducted by Demeke et al, in Arba Minch zuria woreda. As we have not assessed the viability of the gametocytes, mere detection of presence was argued for its sterilizing effect [31]. Also, the sizable anemia prevalence in our participants, 30% on enrolment, might have contributed to the delayed clearance observed. Additionally, the high initial parasite load in 56.1%, over half of our participants (geometric mean of 13,513 parasites/μL) could contribute to delayed clearance and increased risks of treatment failure.

The association between drug efficacy and parasite biomass is multifactorial and can be influenced by various factors such as hemoglobin level, host immune response, drug resistance, and individual variations. The higher parasite biomass during drug initiation can affect the rate of parasite clearance and increase risks of tolerance, which is an important measure of drug efficacy [32]. Although CQ has been officially ceased for more than two decades in Ethiopia for the treatment of *P. falciparum* indicates the presence of indirect CQ pressure in the country, which may impact malaria treatment strategies, and this highlights the need for continued monitoring of antimalarial resistance in the country [33].

As one of the key elements of CQ resistance, Pfcrt-76T high prevalence has been observed in a previous study in Ethiopia, particularly in Adama, Metehara, and Olenchiti, where it was found in 95.7%, 92.5%, and 84.5% of isolates, respectively [33]. The Sudanese study that sequenced samples collected from 2016 to 2020 close to the Ethiopian border detected [34] *PfK13-*R622*I* mutation in 53.8% while in our study, the proportion was not assessed.

Strikingly, the efficacy was 93% in Sudan and 91.3% in Ethiopia, when the threshold for drug change is 90% or less as per WHO [15]. Moreover, the rapid increase in the proportion of detection of the *PfK13*-R662I in the region and broader is alarming. The Current study in five regions of Ethiopia reveals the proportion of R622I rose to 15.7% with additional previously validated by WHO as artemisinin resistance, particularly P675V in Sudanese refugee clinic in Gambella region of Ethiopia [35].

In our study, although the AL efficacy remains near the range of the WHO threshold, we suggest that the combination of a high proportion of MOI and the observed high prevalence of residual submicroscopic parasitemia after ACT treatment may contribute to the decline in efficacy.

As we found high MOI, which aligned with other single nucleotide point mutations, can lead to drug resistance and treatment failure, as different parasite genotypes may have varying susceptibility to antimalarial drugs [36]. Following this, the malaria elimination and control program may be hindered due to the high prevalence of MOI with high recombination rates, harboring multiple genetically distinct parasite clones in endemic settings leading to diverse parasite clone isolates, contributing to high production of gametocytes, rapid emergence, and distribution of drug-resistant *P. falciparum* parasite strains that contribute lowering efficacy [3739]. Notably, the PfK13-R622I mutation was initially identified in 2016 in northern Ethiopia, with 2.4% prevalence, which rose to 9.5% in 2018 [40], and a higher proportion (9.5%) was detected in the samples collected in 2018 from the same area [41] and rose 15.7% sample pooled from five regions of Ethiopia. A similar trend was observed in Eritrea, where the mutation increased from 8.6% in 2016 to 21.0% in 2019 [42]. This increasing trend in the proportion of P. falciparum strains harboring the PfK13-R622I mutation over a short period calls for an explicit understanding of the comparative advantage the mutation confers.

The combination of AL plus SLD PQs showed a better effect in reducing the gametocyte carriage and successful clearance of asexual parasites [31]. On the other hand, the duration of gametocyte clearance was long in our study; of the 10 participants with detectable gametocytes, 1 remained gametocytemic at day 3, and complete clearance was achieved by day 7. Given that the efficacy observed is alarmingly close to the WHO threshold of 90%, our study suggests that the combination of high MOI, high density of pretreatment parasite, and residual parasitemia may contribute to declining efficacy rates.

### Study limitations

The absence of Pfkelch13 and other resistance marker sequencing hinders direct attribution of the 91.3% efficacy to specific genetic variants. Drug level measurements were not performed, preventing assessment of adherence or pharmacokinetic factors. The study was not powered to evaluate gametocyte clearance, limiting conclusions about PQ’s transmission-blocking efficacy.

## Conclusions

The PCR-corrected efficacy of AL+PQ, marginally above the WHO’s threshold, indicates a concerning decline in AL’s effectiveness in this high transmission setting. High MOI, pretreatment parasite density, and potential residual submicroscopic parasitemia may contribute to this reduced efficacy. Continuous monitoring using advanced molecular approaches, such as next-generation sequencing (NGS), is essential to detect resistance-associated mutations early and inform Ethiopia’s malaria treatment policies. Strengthened surveillance is critical to sustain malaria control progress in the study area and beyond.

## Data Availability

All necessary data are all contained within the manuscript and will be shared without limitation upon request specifically

## Acknowledgments

The authors would like to thank Benishangul Gumuz regional and district health offices, study participants, Bambasi health center laboratory technicians, study nurses, and Armauer Hansen Research Institute drivers for their invaluable support and facilitation during the study.

## Author contributions

**Conceptualization:** Jimma Dinsa Deressa, Sinkinesh Wolde Behaksra, Fitsum Girma Tadesse, & Endalamaw Gadisa

**Sample collection and Project Management**: Jimma Dinsa Deressa, Sinkinesh Wolde Behaksra, Kidist Woldekidan & Girma Shumie

**Data Curation**: Jimma Dinsa Deressa, Fikregabrail Aberra Kassa

**Formal Data Analysis**: Jimma Dinsa Deressa, Legesse Alamerie Ejigu, Tiffany Huwe, Eshetu Molla, Yehenew Asmamaw, Bethlehem Adnew, Alemayehu Letebo, & Fikregabrail Aberra Kassa

**Investigations:** Jimma Dinsa Deressa, Dawit Hailu Alemayehu, Tamrayehu Seyoum, Elias Bekele, Sinkinesh Wolde, Tiffany Huwe, Alemayehu Letebo & Migbaru Keffale

**Methodology:** Jimma Dinsa Deressa, Yehenew Asmamaw, Eshetu Molla, Alemayehu Letebo Albejo, Migbaru Keffale, Sinkinesh Wolde Behaksra, Fitsum Girma Tadesse, & Endalamaw Gadisa

**Supervision:** Bayissa Chala, Yehenew Asmamaw, Fitsum Girma Tadesse, and Endalamaw Gadisa **Original Draft:** Jimma Dinsa Deressa, Yehenew Asmamaw, Sinkinesh Wolde Behaksra, Gudissa Assefa, Endalamaw Gadisa

**Review and Editing:** Jimma Dinsa Deressa, Bayissa Chala, Eshetu Molla, Cristian Koepfli, Gudissa Assefa, Fitsum Girma Tadesse, & Endalamaw Gadisa

## Data and code availability

The data code and metadata will be available upon request

## Ethical consideration

The AHRI/ALERT ethics committee (Po/23/19) approved this study. Additionally, this in vivo efficacy trial study was registered under Pan-African clinical trials registry (PACTR) with unique identification number of PACTR202509595696440.Written informed consent was obtained from all participants or their parents/legal guardians, with additional assent secured from participants aged 11 to 17 years.

